# A Comparison of Contemporary versus Older Studies of Aspirin for Primary Prevention

**DOI:** 10.1101/19004267

**Authors:** Frank Moriarty, Mark H. Ebell

## Abstract

**Objective:** This study compares the benefits and harms of aspirin for primary prevention before and after widespread use of statins and colorectal cancer screening.

**Methods:** We compared studies of aspirin for primary prevention that recruited patients from 2005 onward with previous individual patient meta-analyses that recruited patients from 1978 to 2002. Data for contemporary studies were synthesized using random-effects models. We report vascular (major adverse cardiovascular events [MACE], myocardial infarction [MI], stroke), bleeding, cancer, and mortality outcomes.

**Results:** The IPD analyses of older studies included 95,456 patients for CV prevention and 25,270 for cancer mortality, while the four newer studies had 61,604 patients. Relative risks for vascular outcomes for older vs newer studies follow: MACE: 0.89 (95% CI 0.83-0.95) vs 0.93 (0.86-0.99); fatal hemorrhagic stroke: 1.73 (1.11-2.72) vs 1.06 (0.66-1.70); any ischemic stroke: 0.86 (0.74-1.00) vs 0.86 (0.75-0.98); any MI: 0.84 (0.77-0.92) vs 0.88 (0.77-1.00); and non-fatal MI: 0.79 (0.71-0.88) vs 0.94 (0.83-1.08). Cancer death was not significantly decreased in newer studies (RR 1.11, 0.92-1.34). Major hemorrhage was significantly increased for both older and newer studies (RR 1.48, 95% CI 1.25-1.76 vs 1.37, 95% CI 1.24-1.53). There was no effect in either group on all-cause mortality, cardiovascular mortality, fatal stroke, or fatal MI.

**Conclusions:** In the modern era characterized by widespread statin use and cancer screening, aspirin does not reduce the risk of non-fatal MI or cancer death. There are no mortality benefits and a significant risk of major hemorrhage. Aspirin should no longer be recommended for primary prevention.

**Summary of current evidence and what this study adds:** *What is already known about this subject?:*

- The cumulative evidence for aspirin suggests a role in the primary prevention of cardiovascular disease, and in reducing cancer incidence and mortality.
- However most of the trials of aspirin for primary prevention were set in Europe and the United States and recruited patients prior to the year 2000.
- The benefits and harms of aspirin should be considered separately in studies performed in the eras before and after widespread use of statins and colorectal cancer screening.

*What does this study add?:*

- This study provides the most detailed summary to date of cardiac, stroke, bleeding, mortality and cancer outcomes to date in the literature.
- In trials of aspirin for primary prevention from 2005 onwards, aspirin reduced major adverse cardiovascular events but significantly increased the risk of bleeding, with no benefit for mortality or,
- Unlike older studies, there was no reduction in cancer mortality and non-fatal myocardial infarction.

*How does this impact on clinical practice?:*

- Our study suggests aspirin should not be recommended for primary prevention in the modern era.

## Introduction

The first systematic review suggesting a role for aspirin in cardiovascular prevention was published over 30 years ago, and reported relative risk reductions of approximately 15% for cardiovascular mortality and 30% for non-fatal myocardial infarction (MI) and non-fatal stroke in patients with known vascular disease.^1^ However, a subsequent analysis of aspirin for primary prevention by the same group found no reduction in the risk of cardiovascular death or non-ischemic stroke. While the relative risk reduction for non-fatal MI was similar to that in secondary prevention (29% vs 35%), the absolute risk reduction was much smaller for primary prevention (0.5% primary vs 1.4% secondary), resulting in a much larger number needed to treat (NNT) to prevent one non-fatal MI.^2^

More recently, an individual patient data (IPD) meta-analysis of 8 trials with 25,570 patients found a reduction in cancer deaths for patients taking aspirin for 5 or more years,^3^ and another study reported a reduction in the incidence of colorectal cancer in aspirin users.^4^ However, systematic reviews have also consistently found that in both primary and secondary cardiovascular prevention populations, aspirin users had a significantly increased risk of major intracranial and extracranial bleeding.^5^

Most of the trials of aspirin for primary prevention were set in Europe and the United States and recruited patients prior to the year 2000,^5^ a period before statins were in wide use.^6,7^ For example, the Women’s Health Study reported only a 3% rate of statin use at baseline for women recruited between 1992 and 1995.^8^ This was also an era when many countries did not have high rates of screening for colorectal cancer.^9,10^ Statins have been shown to be effective for primary cardiovascular prevention,^11^ and screening has also been shown to reduce both the incidence and mortality due to colorectal cancer.^12^ Four recent trials, done during an era when both use of statins and colorectal cancer screening are much more widespread, found similar or even increased harms, but fewer benefits for aspirin as primary prevention.^13,14,15,16,17,18^ Considering the contemporary benefits and harms in this context is important, particularly for family physicians who consider primary prevention with aspirin for their patients.

A recent meta-analysis added the results of these newer trials to previously published studies and concluded that the overall analysis still favored aspirin for primary prevention of major cardiovascular events, reporting an absolute risk reduction of 0.38%,^19^ similar to that of the older trials. ^5,20^ However, we would argue that it is important to consider separately studies performed in the eras before and after widespread use of statins and colorectal cancer screening, as the contemporary studies more closely reflect the context of modern primary care practice. That is the goal of this meta-analysis.

## Methods

Our study was a limited meta-analysis of the four most recent large trials of aspirin for primary prevention (performed since 2005), and we then compared those results with a previously published individual patient data meta-analysis.

### Study inclusion

This meta-analysis included published studies of randomized controlled trials of aspirin compared to placebo for primary prevention of cardiovascular disease. The population of interest were adults with no history of vascular disease, and specifically those recruited from 2005 onwards, characterizing the modern era of widespread statin prescribing and colorectal cancer screening. This is based on studies showing a large increase in the prevalence of statin use following publication of the 2001 National Cholesterol Education Program Adult Treatment Protocol III guidelines.^6,21^ We also included a previously published meta-analysis from the Antithrombotic Trialists (ATT) Collaboration.^3,5^ This used individual patient data from aspirin primary prevention trials that recruited participants between 1978 and 1998.

### Data items

We extracted data from the modern era trials relating to study characteristics (recruitment year, duration of follow-up, number of participants), baseline participant characteristics (age, sex, body mass index greater than or equal to 25kg/m^2^, current smoking, statin use, and diagnosis of type 2 diabetes mellitus) and efficacy and safety outcomes. We extracted outcome data as the number of participants with each outcome of the following: major adverse cardiovascular event (a composite of cardiovascular death, non-fatal stroke, or non-fatal myocardial infraction), mortality (all-cause, cardiovascular, cancer), cancer incidence, bleeding (major hemorrhage, intracranial hemorrhage), stroke (any, ischemic, and hemorrhagic, each subdivided into any, fatal, and non-fatal), and myocardial infraction (any, fatal, and non-fatal). We also extracted pooled outcome data from the ATT collaboration meta-analysis where available.^5^ The ATT collaboration adjusted the number of events and participants for one study to account for a 2:1 allocation ratio of aspirin vs control, and we used the adjusted numbers in our analysis.

### Data extraction

We developed a data extraction sheet, which we pilot tested and modified for final extraction. We extracted data independently in duplicate. Any discrepancies were discussed and resolved by consensus. We used the intention-to-treat population. In cases where conflicting data were reported within the same publication, we used the most detailed source e.g. supplementary tables provided in an appendix. Where items were not reported in the primary trial publication, we searched and extracted data from other sources where available, including other publications arising from the same trial or from clinical trial registries (e.g. www.clinicaltrials.gov). For any items that were still missing, we contacted corresponding authors of the primary trial publication and further publications where available to request these data.

### Analysis

We summarized trial and participant characteristics for the included studies. The primary measure of treatment effect was the relative risk of each outcome in the aspirin group compared to the placebo group. For statin era trials, relative risks with 95% confidence intervals were computed using random-effects models (taking the estimate of heterogeneity from the Mantel-Haenszel model). We plotted study-specific and summary effect measures with confidence intervals on forest plots to visually assess heterogeneity, and also quantified heterogeneity using the I^2^ statistic. We used the summary estimate of the relative risk reduction or increase and the pooled estimate of the outcome in the control group to calculate the risk of that outcome in the aspirin group. This was in turn used to compute the absolute risk reduction or increase for each outcome as well as the number needed to treat for benefit or harm. We tabulated summary effect measures from this meta-analysis and the ATT collaboration meta-analysis for comparisons. Analyses were performed using Stata 15.1 (StataCorp, College Station, TX, USA) with statistical significance assumed at p<0.05. Extracted data and analytical code from this study are available at www.doi.org/10.5281/zenodo.3149365.

## Results

### Included studies

We identified 6 reports of 4 large randomized controlled trials that recruited a total of 60,604 patients from 2005 onwards: Aspirin in Reducing Events in the Elderly (ASPREE), Aspirin to Reduce Risk of Initial Vascular Events (ARRIVE), A Study of Cardiovascular Events in Diabetes (ASCEND), and the Japanese Primary Prevention Project (JPPP).^13,14,15,16,17,18^ While a study by Fowkes and colleagues was published in 2010, it was excluded because it actually recruited patients between 1998 and 2001.^22^ Two publications were used to ascertain outcomes in older trials, an individual patient data meta-analysis of 6 studies of 95,456 patients recruited from 1978 to 1998 for vascular and bleeding outcomes^5^ and a meta-analysis of 8 studies of the effect of aspirin cancer mortality with 25,570 patients that recruited patients from 1978 to 2002.^3^ Key patient and study characteristics are summarized in Table 1. We were able to abstract data for most outcomes from all four of the contemporary trials, although we were unable to obtain data regarding cancer death and some stroke subtypes for the ARRIVE trial (the full data table and Stata code are available in the online Appendix).

**Table 1.** Baseline characteristics of patients in included studies

| Study (number of participants) | Recruitment Years | Years Follow-Up | Mean Age (years) | BMI > 25 kg/m<sup>2</sup> | Mean BMI (kg/m<sup>2</sup>) | Female (%) | Current Smoker (%) | Statin Use (%) | T2DM (%) |
| --- | --- | --- | --- | --- | --- | --- | --- | --- | --- |
| **Older meta-analysis** |  |  |  |  |  |  |  |  |  |
| ATT, 2009<br>(n=95,456) | 1978 – 1998 | 3.6 – 10 years | 56 | NR | 26.3 | 54% | 16% | NR | 4.0% |
| ATT, 2011<br>(n=25,570) | 1978 – 2002 | 4.2 – 6.9 years<br>(median) | 59.8 | NR | NR | 29.4% | 35.1% | NR | NR |
| **New Trials** |  |  |  |  |  |  |  |  |  |
| JPPP, 2014<br>(n=14,464) | 2005-2007 | 5.0 years<br>(median) | 70.5 | 5248<br>(35.3%) | 24.2 | 8341<br>(57.7%) | 1893 (13.1%) | NR ** | 4903<br>(33.9%) |
| ASCEND, 2018<br>(n=14,480) | 2005-2011 | 7.4 years<br>(mean) | 63.3 | 12730<br>(82.2%) | 30.7 | 5796<br>(37.4%) | 1279 (8.3%) | 11,653<br>(75.3%) | 14,569<br>(94.1%) |
| ARRIVE, 2018<br>(n=12,546) | 2007-2013 | 5.1 years<br>(median) | 63.9 | 9854<br>(78.5%) | 28.4 | 3708<br>(29.6%) | 3594 (28.6%) | 5455<br>(43.5%) | 0 (0.0%) |
| ASPREE, 2018<br>(n=19,114) | 2010-2014 | 4.7 years<br>(median) | 74* | 14157<br>(74.1%) | 28.1 | 10783<br>(56.4%) | 735 (3.8%) | 6470<br>(33.8%) | 2057<br>(10.8%) |
| **Total (new trials):** | 2005 to 2014 | 4.7 – 7.4 years | 68.5 | 69.3% | 27.9 | 47.2% | 12.4% | 51.1% + | 35.5% |

\* Median age
\*\* Not reported, but 72% described as having comorbid hyperlipidemia at enrollment.
+ Excluding JPPP. If included with the assumption that 80% of patients with hyperlipidemia on enrollment were taking a statin, prevalence of statin use is 52.7%
BMI = body mass index, T2DM = type 2 diabetes mellitus, NR = not reported

**Table 2.** Summary of benefits and harms from older and newer studies of aspirin for primary prevention (bold face = statistically significant at p < 0.05).

| Outcome | Older Studies<sup>3,5</sup> | Most Recent Studies<sup>13,14,15,16,17,18</sup> |
| --- | --- | --- |
| **Major Adverse Cardiovascular Events \*** | **0.89 (0.83, 0.95)** | **0.93 (0.86, 0.99)** |
| **Mortality Outcomes** |  |  |
| All-cause mortality | 0.95 (0.89, 1.01) | 1.01 (0.92, 1.12) |
| Cardiovascular mortality | 0.97 (0.87, 1.09) | 0.92 (0.81, 1.06) |
| Fatal myocardial infarction | 0.95 (0.82, 1.09) | 0.84 (0.67, 1.06) |
| Fatal stroke | 1.21 (0.93, 1.59) | 1.02 (0.71, 1.45) |
| **Bleeding Outcomes** |  |  |
| Intracranial hemorrhage | NR | **1.44 (1.16, 1.80)** |
| Major hemorrhage | **1.53 (1.29, 1.81)** | **1.37 (1.24, 1.53)** |
| Stroke (any hemorrhagic) | 1.30 (0.99, 1.72) | 1.23 (0.92, 1.64) |
| **Cancer Outcomes** |  |  |
| Cancer death | **0.79 (0.68, 0.92)\*\*** | 1.11 (0.92, 1.34) |
| Cancer incidence | NR | 1.06 (0.99, 1.14) |
| **Stroke Outcomes** |  |  |
| Stroke (any) | 0.96 (0.86, 1.07) | 0.96 (0.86, 1.07) |
| Stroke (any fatal) | 1.21 (0.93, 1.59) | 1.02 (0.71, 1.45) |
| Stroke (any non-fatal) | 0.93 (0.83, 1.04) | 0.93 (0.82, 1.05) |
| Stroke (any hemorrhagic) | 1.30 (0.99, 1.72) | 1.23 (0.92, 1.64) |
| Stroke (fatal hemorrhagic) | **1.73 (1.11, 2.72)** | 1.06 (0.66, 1.70) |
| Stroke (non-fatal hemorrhagic) | 1.09 (0.76, 1.55) | 1.39 (0.80, 2.42) |
| Stroke (any ischemic) | 0.86 (0.74, 1.00) | **0.86 (0.75, 0.98)** |
| Stroke (fatal ischemic) | 0.83 (0.48, 1.42) | 0.98 (0.56, 1.72) |
| Stroke (non-fatal ischemic) | 0.87 (0.74, 1.01) | 0.88 (0.77, 1.00) |
| **Myocardial Infarction Outcomes** |  |  |
| Any myocardial infarction | 0.94 (0.88, 1.03) | 0.88 (0.77, 1.00) |
| Fatal myocardial infarction | 0.95 (0.82, 1.09) | 0.84 (0.67, 1.06) |
| Non-fatal myocardial infarction | **0.79 (0.71, 0.88)** | 0.90 (0.76, 1.06) |

\* Cardiovascular death, non-fatal myocardial infarction, and non-fatal stroke
\*\* Data for cancer death from meta-analysis of eight studies;<sup>3</sup> all other data from individual patient meta-analysis of 6 studies.<sup>5</sup>

**Table 3.** Estimated events per 1000 for selected outcomes in groups treated with aspirin and controls, using summary estimates of relative risk increase or reduction from statin era studies applied to the prevalence of outcomes in the control groups.

| Outcome | Rate in control group | Rate in aspirin group based on summary estimate of RR (95% confidence interval) | Absolute risk increase or reduction (95% confidence interval) | Number needed to treat (NNT) or harm (NNH) over study duration (95% confidence interval) | Events / 1000 persons treated for ~ 5 years |  |
| --- | --- | --- | --- | --- | --- | --- |
|  |  |  |  |  | Aspirin | Control |
| Major adverse cardiovascular events * | 1468/30,849 = 4.76% | 4.43% (4.09%, 4.71%) | 0.33% (0.05%, 0.67%) | 303 (149, 2000) | 44 | 48 |
| Any ischemic stroke | 560/30,849 = 1.81% | 1.56% (1.36%, 1.78%) | ARR = 0.25% (0.03%, 0.45%) | NNT = 400 (222, 3333) | 16 | 18 |
| Intracranial hemorrhage | 132/24,573 = 0.54% | 0.78% (0.63%, 0.97%) | ARI = 0.24% (0.09%, 0.43%) | NNH = 417 (233, 1111) | 8 | 5 |
| Major hemorrhage | 585/30,849 = 1.90% | 2.60% (2.36%, 2.91%) | ARI = 0.70% (0.46%, 1.01%) | NNH = 143 (99, 217) | 26 | 19 |

\* Cardiovascular death, non-fatal myocardial infarction, and non-fatal stroke.

The patient populations differed considerably between older and newer trials. Compared with patients in the individual patient data meta-analysis for cardiovascular outcomes,^5^ patients in the 4 newest trials were older (68.5 vs 56 years), were somewhat less likely to smoke (12% vs 16%) and were much more likely to have type 2 diabetes mellitus (T2DM, 35.5% vs 4.0%). Compared with patients in the meta-analysis of older studies for the association between aspirin and cancer,^3^ patients in the newer studies were older (69 vs 60 years), more likely to be female (47% vs 29%) and were much less likely to be smokers (12% vs 35%). Broadly, the participants in the newer trials appear to be consistent with a contemporary primary care population at increased risk for vascular disease that would be considered for aspirin as primary prevention.

### Clinical benefits and harms of aspirin

There was no difference between aspirin and placebo groups in any of the mortality outcomes, including all-cause mortality and cardiovascular mortality, for both older and newer studies. Both older and newer studies found a significant reduction in major adverse cardiovascular events, a composite of cardiovascular death, non-fatal myocardial infarction (MI) and non-fatal stroke. The reduction was smaller in newer studies (RR 0.93 vs 0.89) but remained statistically significant. See Figure 1 for a forest plot of these outcomes in the newer studies, and eFigure 1 for these outcomes in the older trials.

**Figure 1.**
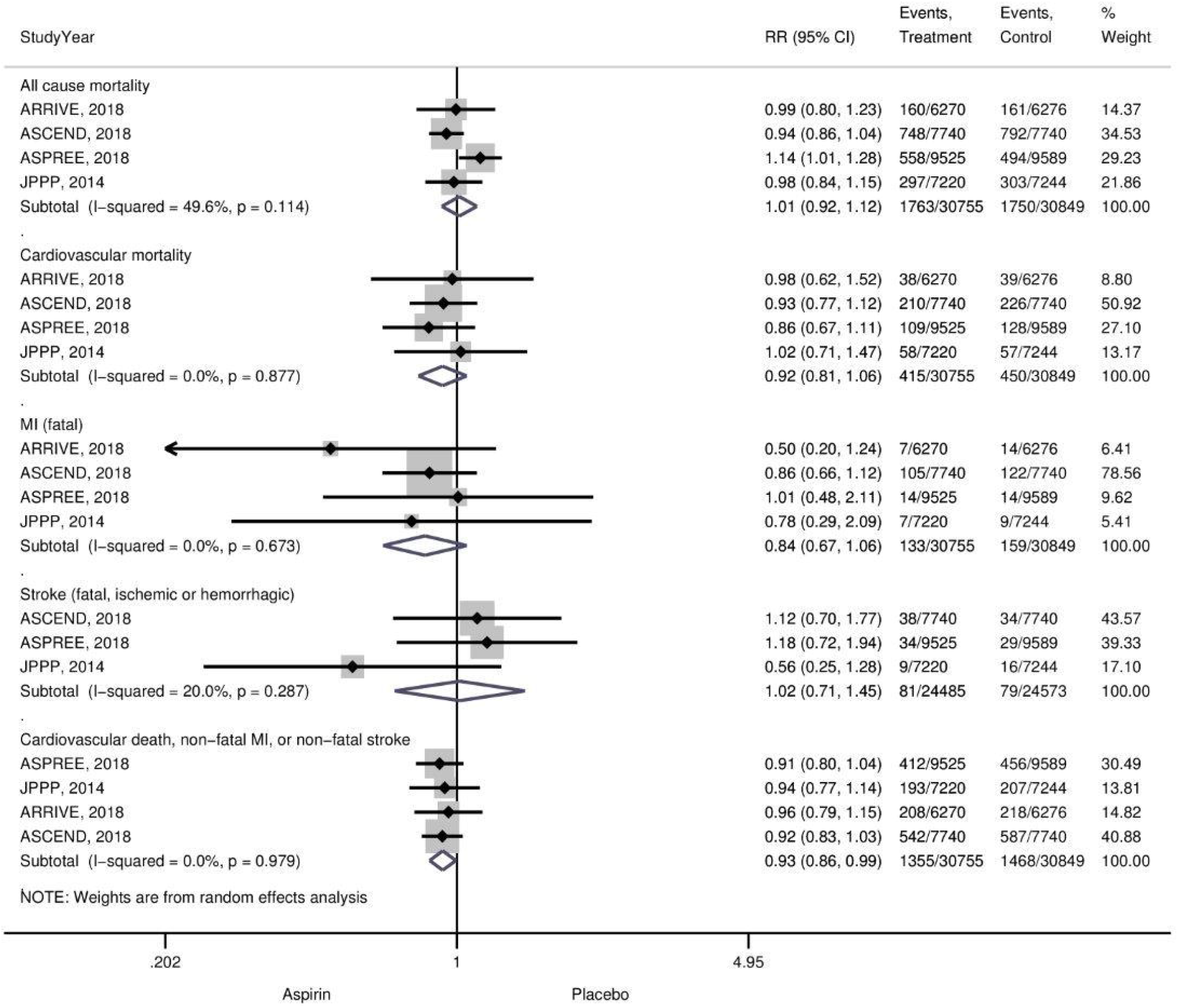
Forest plot of mortality outcomes and major adverse cardiovascular events for 4 newer studies of aspirin for primary prevention.

There was a clinically and statistically significant increase in the risk of major hemorrhage for both older and newer studies, as well as an increase in the risk of intracranial hemorrhage. Regarding cancer, while the meta-analysis of eight older studies reported a reduction in the risk of cancer death (RR 0.79, 95% CI 0.68-0.92), this was not seen in the newer trials (RR 1.11, 95% CI 0.92-1.34). Cancer incidence was not reported by the older meta-analysis and was higher although significantly in the newer studies (RR 1.06, 95% CI 0.99-1.14). See Figure 2 for a forest plot of these outcomes in the newer studies, and eFigure 1 for these outcomes in the older trials.

**Figure 2.**
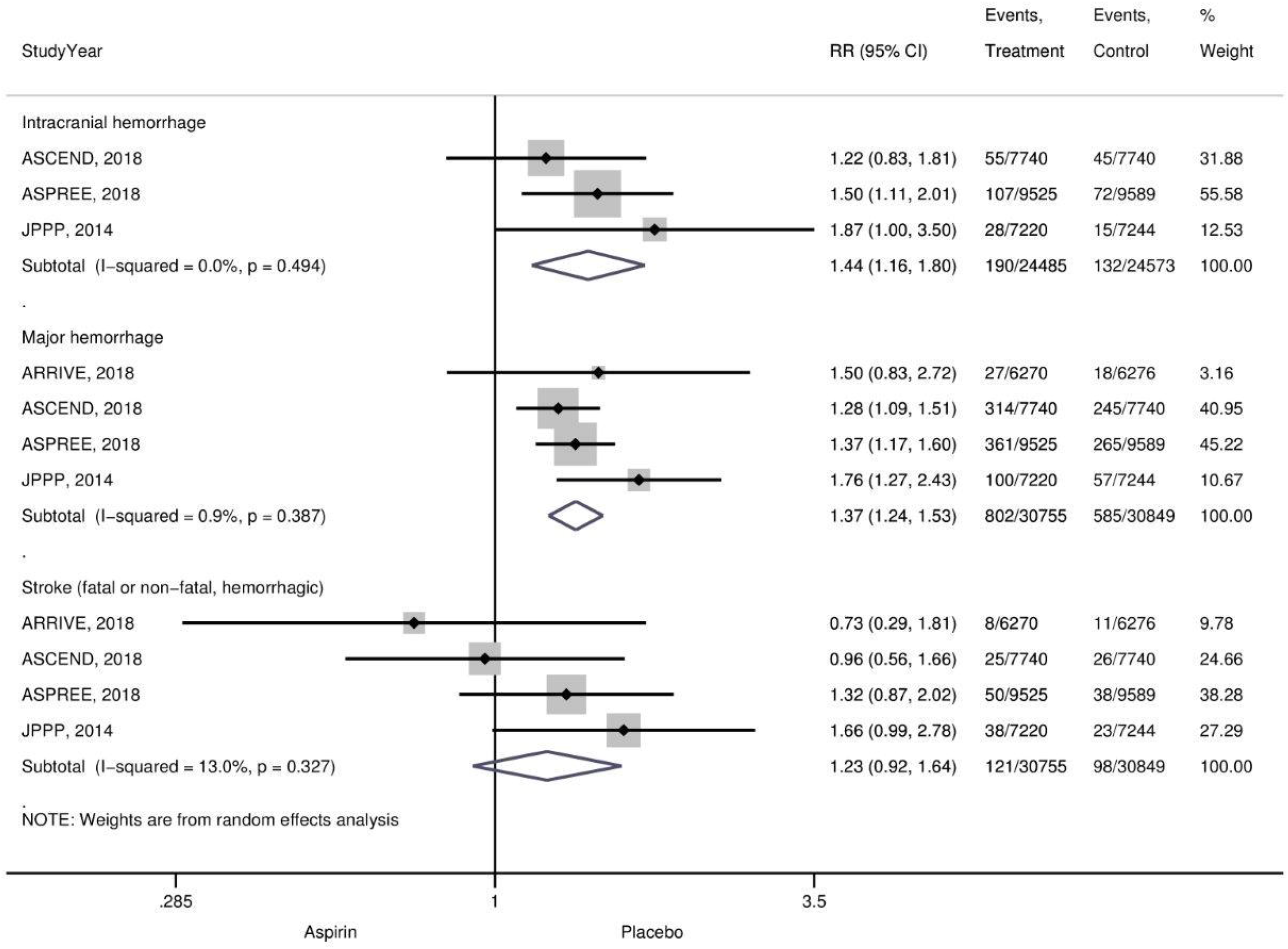
Forest plot of bleeding outcomes for 4 newer studies of aspirin for primary prevention.

There was little difference between outcomes for various stroke subgroups by fatal vs non-fatal vs any stroke and ischemic vs hemorrhagic vs any type of stroke. An increase in the risk of fatal hemorrhagic stroke in older studies was not seen in the newer studies, and the likelihood of any ischemic stroke was identical in magnitude to that seen in older studies (RR 0.86 vs 0.86) but was now statistically significant.

Regarding myocardial infarction, older studies found a significant benefit to aspirin with fewer non-fatal MIs (RR 0.79, 95% CI 0.71-0.88), but there was no reduction in fatal MIs. In newer studies, there was no significant reduction in the likelihood of fatal or non-fatal MI. Forest plots for stroke, myocardial infarction and cancer outcomes for all newer studies are summarized in the Appendix available online (Appendix Figures 2 – 4).

The lower relative risk of major adverse cardiovascular events equates to an absolute risk reduction of 0.33% (95% CI 0.05%-0.67%) and for the outcome of any ischemic stroke, it corresponds to an absolute risk reduction of 0.25% (95% CI 0.03%-0.45%). In relation to harms, aspirin was associated with absolute risk increases for intracranial hemorrhage and major hemorrhage of 0.24% (0.09%-0.43%) and 0.70% (0.46%-1.01%) respectively.

## Discussion

Like older studies, recent trials of aspirin for primary prevention found no mortality benefit and a significant increase in the risk of major hemorrhage. As with older studies, there was a lower risk of the composite of cardiovascular death, non-fatal MI, and non-fatal stroke in newer studies, which corresponds to a number needed to treat to prevent one event over approximately 5 years of 263 (95% CI 149-2000). However, newer studies also failed to find evidence for two important potential benefits of aspirin: a reduced risk of cancer mortality and a reduced risk of non-fatal MI. Based on this meta-analysis, we estimate that for every 1000 persons who take aspirin instead of placebo for 5 years there will be 4 fewer major cardiovascular events (including 2 fewer ischemic strokes), 3 more intracranial hemorrhages and 7 additional episodes of major hemorrhage with no effect on mortality.

There are a number of potential explanations for the failure of aspirin to reduce the risk of cancer death in newer studies. Patients in older studies were much less likely to have undergone regular cancer screening. For example, the US Medicare program did not pay for colorectal cancer screening until 1998,^10^ mammography was not in widespread use prior to the late 1980’s,^23^ and colorectal cancer screening programs largely started in the late 1990’s or 2000’s in the countries that have adopted it.^9,10^ Cancer treatment has also improved, and overall rates of cancer survival have increased over the past 30 years.^24,25^ Both of these may serve to dampen any potential benefit of aspirin for cancer prevention, by reducing the overall risk of cancer death, and therefore the absolute benefit possible. This is especially true for colorectal cancer screening, which reduces incidence as well as mortality.^26^ In fact, the recent ASPREE trial found a significant increase in the risk of cancer death with aspirin (3.1% vs 2.3%, HR 1.31, 95% CI 1.10-1.56).^13^

A limitation of newer trials with regards to the effect on cancer mortality is their relatively short duration of approximately 5 years for three studies, with the fourth following patients for a mean of 7.4 years (ASCEND). This may not be long enough to see a benefit in terms of cancer mortality, which did not appear until after 5 years of follow-up in the individual patient meta-analysis performed by Rothwell and colleagues. On the other hand, there was also a trend toward increased cancer incidence in the newer trials (RR 1.06, 95% CI 0.99 - 1.14). Another limitation of our analysis was differences in the inclusion criteria for some outcomes, most notably major hemorrhage, resulting in a broad range of absolute risks for that outcome in the newer studies (0.4% to 3.6%). However, the relative risk increases were consistent (RR 1.28 to 1.76, i^2^ = 1%, across 4 newer studies). Finally, incomplete reporting of outcomes (especially by the ARRIVE trial) limited the sample to 3 of the newer studies for some outcomes.

Management of cardiovascular disease has also changed considerably from the 1980’s and 1990’s to the era after 2005. The publication of the NCEP ATP III guidelines led to large increases in the use of statins, which primarily reduce the risk of non-fatal MI when given as primary prevention. Statins also appear to have an anti-inflammatory effect and have been shown to reduce circulating levels of c-reactive protein^27^, which may also mitigate the effect of aspirin on inflammation. However, evidence from a Mendelian randomization study suggests c-reactive protein may not have a causal role in coronary heart disease.^28^ Developments in the detection of embolic events in the contemporary era also mean that increasingly small ischemic events are defined as myocardial infarction, and thus the contemporary trials may include these in their endpoints.

The equivocal evidence on aspirin’s benefits and risks in primary cardiovascular prevention is reflected in the variable recommendations across different clinical guidelines. The United States Preventive Services Task Force (USPTF) 2017 statement on the topic recommends aspirin for adults aged 50-59 years who have a 10-year CVD risk of ≥ 10% (provided they are not at increased risk for bleeding) and are willing to take aspirin for at least 10 years.^29^ Among those aged 60-69 years, the USPSTF recommends any decision to start aspirin should be an individual one based on each person’s preferences and values. The 10-year minimum duration of therapy is based on older trials showing a reduction in cancer mortality, which we would argue should be re-examined in light of the lack of benefit regarding cancer mortality seen in the newer studies.

Similarly, the 2019 American College of Cardiology/American Heart Association guideline states that aspirin might be considered in selected adults aged 40-70 years with no increased bleeding risk and who have an elevated risk of atherosclerotic cardiovascular disease.^30^ The UK National Institute for Health and Care Excellence (NICE) recommends against the routine prescribing of antiplatelet therapy for primary prevention, but does recommend that aspirin be considered for primary prevention in people at high risk of stroke or MI.^31^ However, this recommendation is caveated with statements that such use is off-label and that the benefits and risks should be discussed with the individual when aspirin is being considered. In contrast, the 2016 European Society of Cardiology (ESC) guidance,^32^ as well as the Scottish Intercollegiate Guidelines Network (SIGN),^33^ do not recommend antiplatelet therapy such as aspirin for people (including those with diabetes mellitus) free from cardiovascular disease, citing increased major hemorrhage risk. Our findings would support these recommendations from the ESC and SIGN.

Mahmoud and colleagues recently published a meta-analysis of aspirin trials for primary cardiovascular prevention.^34^ This included a trial sequential analysis, which analogous to an interim analysis of a single randomized trial, assesses the power of the accumulated evidence to detect or refute an intervention effect. Based on the eleven aspirin primary prevention trials included in this analysis, no benefit for all-cause mortality was detected and it was concluded that further trials to investigate this effect would not be useful.

In conclusion, in a modern era characterized by widespread statin use and population wide cancer screening, aspirin no longer reduces the risk of cancer death or MI when given as primary prevention. Primary care physicians, as those most likely to see patients where primary prevention may be considered, should weigh up and discuss the absolute risks and benefits in this context. A good way to communicate this information to our patients in primary care is using absolute risk: for every 1000 persons who take aspirin instead of placebo for 5 years there will be 4 fewer major cardiovascular events (including 2 fewer ischemic strokes), 3 more intracranial hemorrhages and 7 additional episodes of major hemorrhage with no change in overall or cardiovascular mortality. We therefore conclude that aspirin cannot be recommended for primary prevention in the modern era.

## Data Availability

Extracted data used in this meta-analysis and analysis code are available at www.doi.org/10.5281/zenodo.3149365.

## Abbreviations

ATT: Antithrombotic Trialists
CI: Confidence interval
ESC: European Society of Cardiology
IPD: Individual patient data
MI: Myocardial infarction
NICE: National Institute for Health and Care Excellence
NNT: Number needed to treat
RR: Relative risk
SIGN: Scottish Intercollegiate Guidelines Network
T2DM: Type 2 diabetes mellitus
USPTF: United States Preventive Services Task Force

## Footnotes

### Contributors

MHE conceived and designed the study. MHE and FM extracted the data, MHE conducted the analysis and MHE and FM interpreted the data. MHE and FM drafted the manuscript, and critical revised and approved the final manuscript. The corresponding author attests that all listed authors meet authorship criteria and that no others meeting the criteria have been omitted. MHE is the guarantor.

### Conflict of interest (Intellectual)

All authors have completed the ICMJE uniform disclosure form at www.icmje.org/coi_disclosure.pdf and declare: support for the study as detailed above; no financial relationships with any organisations that might have an interest in the submitted work in the previous three years; Dr. Ebell served as a member of the US Preventive Services Task Force from 2012 to 2015 and voted on the current USPSTF recommendation regarding aspirin for primary prevention that is discussed in this manuscript.

### Ethical approval

Ethical approval for this meta-analysis of published studies was not required.

### Transparency statement

The lead author affirms that the manuscript is an honest, accurate, and transparent account of the study being reported; that no important aspects of the study have been omitted; and that any discrepancies from the study as planned have been explained.

## Appendix Figures

Appendix Figure 1. Forest plot of all outcomes for the individual patient meta-analysis of older studies.

Appendix Figure 2. Forest plot of myocardial infarction outcomes for 4 newer studies of aspirin for primary prevention.

Appendix Figure 3. Forest plot of stroke outcomes for 4 newer studies of aspirin for primary prevention.

Appendix Figure 4. Forest plot of cancer outcomes for 4 newer studies of aspirin for primary prevention

